# Prognostic and predictive factors for primary chemotherapy in locally advanced breast cancer

**DOI:** 10.1101/2020.01.14.20017467

**Authors:** Clara Borges, Daniela Almeida, Margarida Damasceno

## Abstract

**Background:** Complete pathologic response (pCR) to neoadjuvant chemotherapy in breast cancer is associated to a better locoregional disease control and better long-term prognosis. Our purpose is to identify clinical and pathologic characteristics that influence treatment response, relapse risk and overall survival.

**Methods:** We analyzed 341 women with breast cancer submitted to primary chemotherapy between 2004 and 2017. We compared clinical response, pathologic response and conservative surgery rate, among different tumor and patient features. We assessed survival and relapse rate, according to complete pathologic response and histologic characteristics.

**Results:** Among 341 women who underwent neoadjuvant treatment, 34% obtained pCR and 94,4% expressed a pathologic response.

Patients who achived pCR had better overall survival (HR 0,68, CI95% 0,53-0,87, p=0.002) and better progression free survival (HR 0,78, CI95%, 0,61-1,00, p=0,05). pCR was significantly influenced by histologic grade (0% G1, 18,9% G2 vs 41,5% G3, p-value <0,001), HER2 over-expression (56% vs 22,8%, p-value<0,001), cytokeratin 19 expression (38,6% vs 30,8%, p-value 0,047) and cytokeratin 5 (44% vs 32,4%, p-value 0,002).

Instead, presence of bilateral breast cancer (34,7% vs 0%, p-value 0,007) and hormone receptor expression (20,2% vs 54,3%, p-value <0,001) negatively influenced pCR.

Additionally, overall survival was negatively influenced by cytokeratin 19 expression (HR 0,14, CI95% 0,10-0,20, p<0,001) and presence of inflammatory breast cancer (HR 0,26, CI95% 0,11-0,64, p=0.003).

**Conclusions:** Cytokeratin 19 expression, cytokeratin 5 expression, histologic grade, molecular subtypes and bilaterality are independent predictors for pCR to primary chemotherapy.

pCR influences positively overall survival and progression free survival. However, inflammatory carcinoma and cytokeratin 19 expression are independent predictors for a worse prognosis.

## Introduction

Breast cancer is the most common cancer among women, with lifetime risk up to 20%^1^. Locally advanced breast cancer is best managed by multidisciplinary team employing systemic and locoregional therapy. Most patients with locally advanced breast cancer receive neoadjuvant chemotherapy, prior to surgery, in order to induce a local tumor response and enable breast conservation. Pathologic complete response is associated with better long-term prognosis.^2^

A number of tumor and host characteristics have been found useful in predicting the risk of recurrence and death from breast cancer following optimal therapeutic approach.^3^ As the therapeutic options for breast cancer broaden, it is essential to recognize clinical and pathologic features that influence therapeutic response and outcome in order to quantify the risk of progression in each individual patient and tailor treatment accordingly, leading to a more personalized treatment and follow up recommendation. In this retrospective case-control study, we sought to identify the clinical and pathologic variables that influence pathologic response and long-term survival.

## Methods

### Setting and participants

Patients were evaluated at a university tertiary hospital and consented to participate. A total of 341 patients with breast cancer submitted to primary chemotherapy, from 01 January 2004 to 31 August 2017, were included in the study. The study was approved by the local ethics committee.

**Figure 1.**
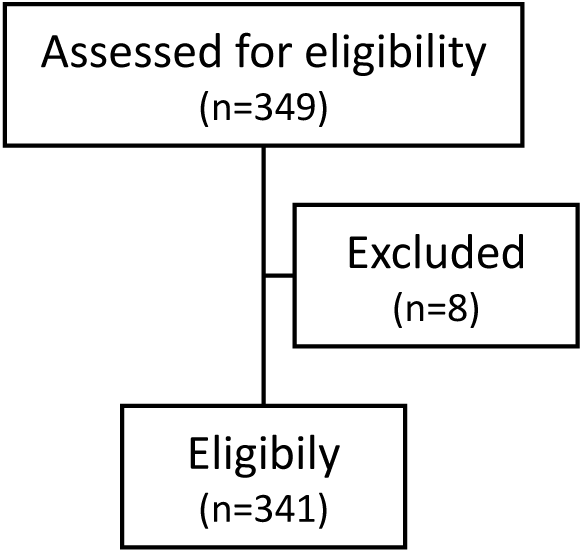
Flow diagram for patient selection.

Eligibility criteria included: female gender, pathologic diagnosis of breast carcinoma, multidisciplinary decision to proceed to neoadjuvant chemotherapy, plus endocrine therapy and/or targeted therapy, according to tumor tissue markers. Eight patients were excluded from the study, due to contraindications to chemotherapy, and underwent primary surgery. The baseline participant characteristics are exhibited in Table 1.

**Table 1.**
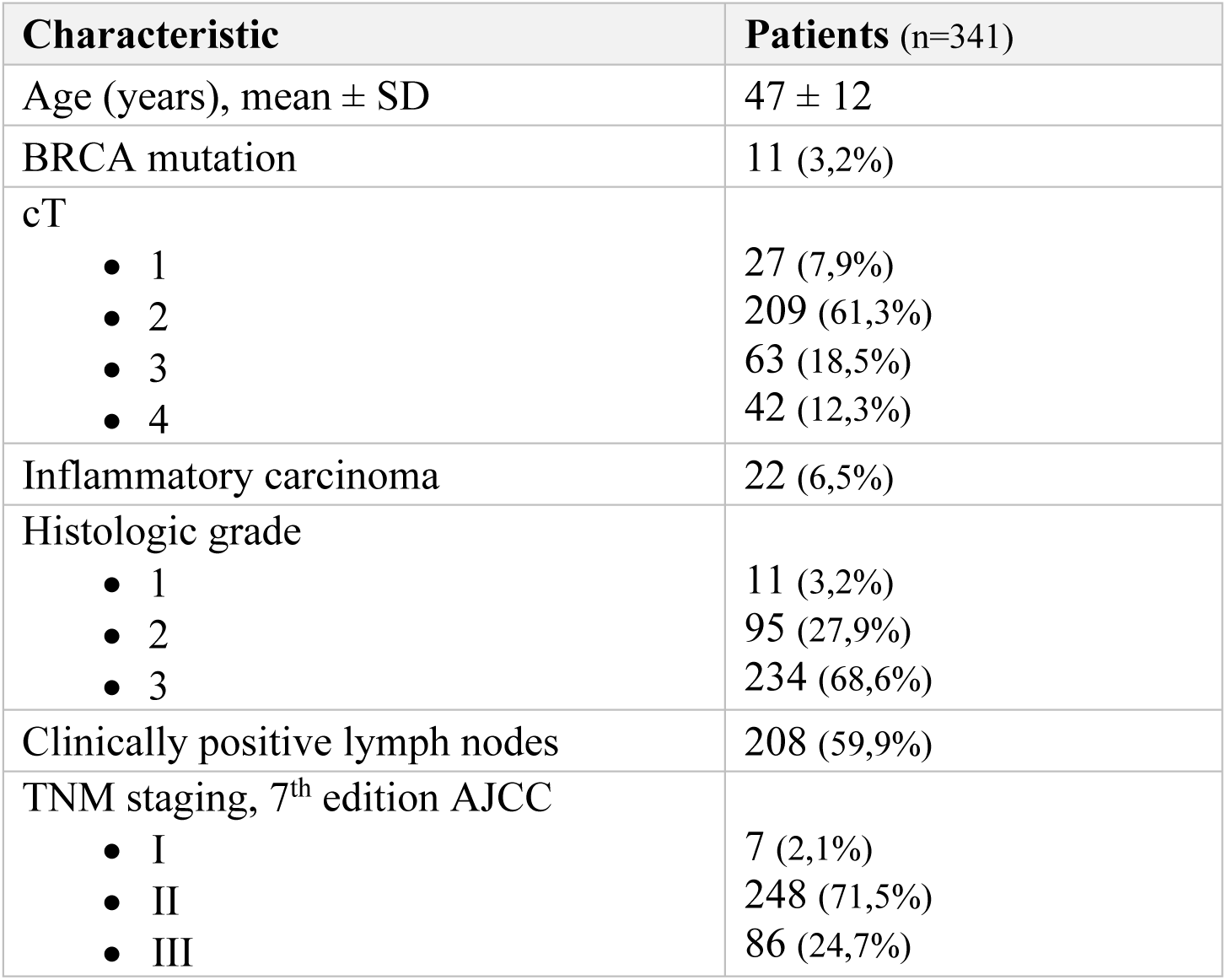
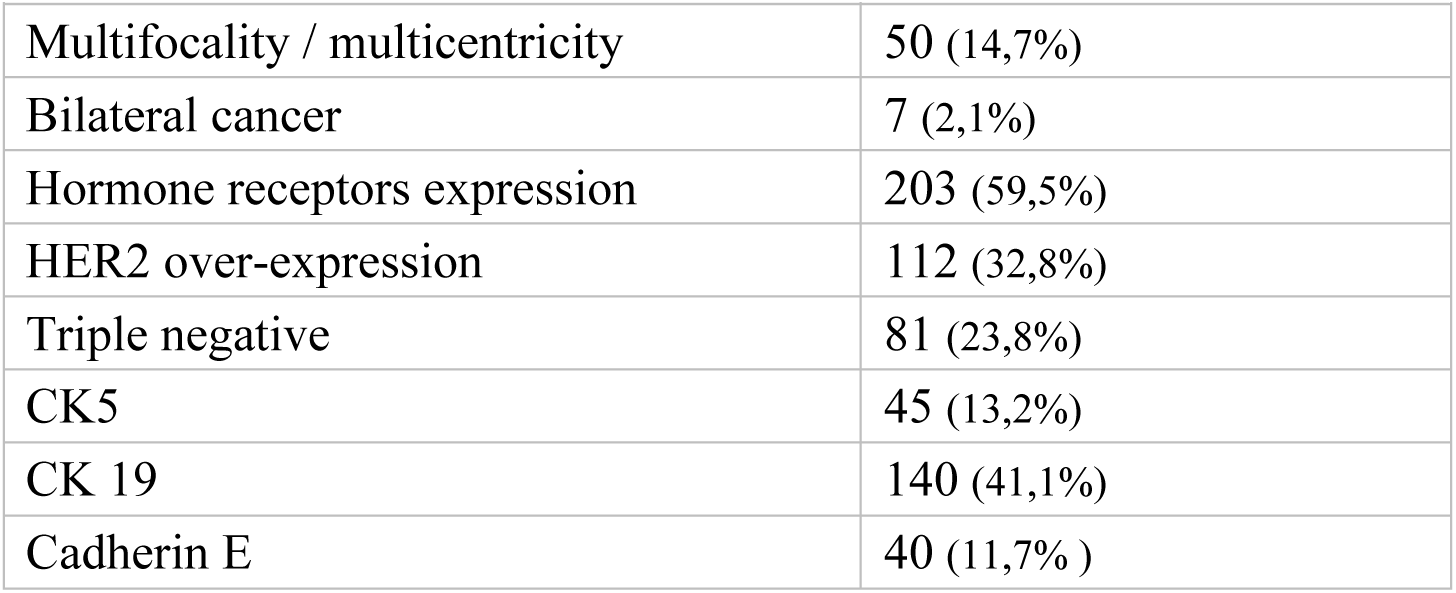
Baseline patients’ characteristics.

### Study design

For the purpose of this retrospective cohort study, data from several clinical variables, such as age, BRCA mutation, size, lymph nodes status, inflammatory carcinoma, multifocality/multicentricity and bilateral breast cancer, was collected. We also examined histologic variables, for example hormonal receptors, HER2 over-expression, histologic grade, presence of cytokeratin 5 (CK5), cytokeratin 19 (CK19) and cadherin E.

### Outcomes and follow up

The primary study end points were complete pathologic response (pCR), progression free survival (PFS) and overall survival (OS). Secondary end points comprised complete clinical response, conservative surgery rate, relapse rate and survival rate.

### Statistical analysis

All comparative analysis was based on chi-square test, Fischer exact test and Kaplan-Meyer plot. P values from chi-square tests were used when the results were consistent otherwise, P values from Fisher’s exact tests were used. Cox regression model was performed, adjusted to age, tumor size, lymph nodes and clinical staging TNM. For statistical analyses, SPSS version 24.0 (SPSS Inc, Chicago) was used. Statistical significance was accepted at values less than 0.05.

## Results

### Clinical and pathologic response analysis

Among 341 women who underwent neoadjuvant chemotherapy followed by surgery, the overall pCR was 34% and pathologic response (complete or partial) was 94,4%.

In our study, pCR rates significantly differed by histologic grade, molecular subtypes, celular markers and presence of bilateral cancer (Table 2). Higher histologic grade, absence of hormone receptors and over-expression of HER2 was associated with better clinical and pathologic response to neoadjuvant chemotherapy. HER2-enriched subtype of breast cancer reported a 64,9% complete pathologic response, in comparison to 10,1% from Luminal A subtype.

**Table 2.** Clinical and pathologic response to neoadjuvant therapy analysis.

| Variables | Clinical and radiological complete response | Conservative surgery rate | Complete pathological response |
| --- | --- | --- | --- |
| • Clinical Variables |  |  |  |
| Age |  |  |  |
| - ≤ 35 y | 13 (24,5%) | 20 (38,5%) | 17 (32,1%) |
| - > 35 y | 90 (31,5%) | 134 (46,5%) | 99 (34,3%) |
|  | p-value 0,224 | p-value 0,282 | p-value 0,413 |
| BRCA mutation |  |  |  |
| - Absence | 99 (30,2%) | 154 (46,8%) | 112 (33,9%) |
| - Presence | 4 (36,4%) | 0 (0%) | 4 (36,4%) |
|  | p-value 0,575 | **p-value 0,002** | p-value 0,828 |
| cT |  |  |  |
| - ≤ 2 | 75 (32,1%) | 132 (56,2%) | 85 (36%) |
| - > 2 | 28 (26,7%) | 22 (21%) | 31 (29,5%) |
|  | p-value 0,152 | **p-value <0,001** | p-value 0,360 |
| Lymph nodes |  |  |  |
| - Clinically positive | 59 (33,1%) | 82 (39,6%) | 68 (32,7%) |
| - Clinically negative | 44 (28,6%) | 72 (54,1%) | 48 (36,1%) |
|  | p-value 0,336 | **p-value 0,009** | p-value 0,572 |
| Inflammatory carcinoma |  |  |  |
| - Absence | 96 (30,3%) | 154 (48,4%) | 108 (33,9%) |
| - Presence | 7 (31,8%) | 0 (0%) | 8 (36,4%) |
|  | p-value 0,725 | **p-value <0,001** | p-value 0,193 |
| Multifocality / multicentricity |  |  |  |
| - Absence | 90 (31%) | 139 (47,9%) | 101 (34,7%) |
| - Presence | 13 (26,5%) | 15 (30%) | 15 (30%) |
|  | p-value 0,435 | **p-value 0,019** | p-value 0,840 |
| Bilateral cancer |  |  |  |
| - Absence | 102 (30,7%) | 153 (45,9%) | 116 (34,7%) |
| - Presence | 1 (14,7%) | 1 (14,3%) | 0 (0%) |
|  | p-value 0,647 | p-value 0,096 | **p-value 0,074** |
| • Pathological Variables |  |  |  |
| Histologic grade |  |  |  |
| - 1 | 0 (0%) | 3 (30%) | 0 (0%) |
| - 2 | 21 (22,1%) | 43 (45,3%) | 18 (18,9%) |
| - 3 | 82 (35,3%) | 108 (46,2%) | 97 (41,5%) |
|  | **p-value 0,030** | p-value 0,634 | **p-value <0,001** |
| Hormone receptors status |  |  |  |
| - Expression | 49 (24,3%) | 85 (42,1%) | 41 (20,2%) |
| - Absence | 54 (39,4%) | 69 (50%) | 75 (54,3%) |
|  | **p-value 0,009** | p-value 0,183 | **p-value <0,001** |
| HER2 status |  |  |  |
| – Over-expression | 46 (41,1%) | 49 (43,8%) | 191 (56%) |
| – Negative | 57 (25,1%) | 105 (46,1%) | 78 (23%) |
|  | **p-value 0,011** | p-value 0,689 | **p-value <0,001** |
| Molecular subtypes |  |  |  |
| – HR+ HER2- | 27 (18,4%) | 58 (39,5%) | 15 (10,1%) |
| – HR+ HER2+ | 22 (40%) | 27 (49,1%) | 26 (47,3%) |
| – HR- HER2+ | 24 (42,1%) | 22 (38,6%) | 37 (64,9%) |
| – Triple negative | 30 (37,5%) | 47 (58%) | 38 (46,9%) |
|  | **p-value 0,004** | **p-value 0,035** | **p-value <0,001** |
| CK 19 |  |  |  |
| – Expression | 51 (37%) | 70 (50%) | 54 (38,6%) |
| – Absence | 52 (25,9%) | 84 (42%) | 62 (30,8%) |
|  | p-value 0,051 | p-value 0,145 | **p-value 0,047** |
| CK5 |  |  |  |
| – Expression | 17 (37,8%) | 26 (57,8%) | 20 (44,4%) |
| – Absence | 86 (29,3%) | 128 (43,4%) | 96 (32,4%) |
|  | p-value 0,136 | **p-value 0,036** | **p-value 0,002** |
| Cadherin E |  |  |  |
| – Expression | 11 (27,5%) | 14 (35%) | 14 (35%) |
| – Absence | 92 (30,8%) | 140 (46,7%) | 102 (33,9%) |
|  | p-value 0,406 | p-value 0,164 | p-value 0,403 |

We observed that CK19 expression was statistically correlated with better clinical and pathologic response to primary chemotherapy, presenting a 38,6% rate of pCR in patients with CK19 positive versus 30,8% CK 19 negative (p-value 0,047). CK5 expression was also related to better pathologic response, displaying a 44,4% rate of pCR versus 32,4% (p-value 0,002).

Conservative surgery was seldom indication in women with large original lesions, inflammatory breast cancer, multifocal/multicentric lesions or clinically-positive lymph nodes. Triple negative neoplasia was more frequently submitted to conservative surgery than other molecular subtypes.

The following factors didn’t influence the complete clinical or pathologic response: younger age, BRCA mutation, inflammatory carcinoma, clinically-positive lymph nodes and cadherin E expression.

### Survival analysis by pCR

In order to determine whether achieving pCR was a prognostic factor in locally advanced breast cancer, we performed survival analysis for 341 women who underwent neoadjuvant chemotherapy. Multivariate analysis (Graphic 1) showed that there was a significant difference in terms of OS and PFS between patients with pCR and those without pCR. Patients who obtained a pCR had a statistically improvement on 5-year OS (82,4% vs 76,3%, p-value <0,0001) and non-statistically significant improvement on 5- year PFS (68,6% vs 54,8%). Thus, the data showed that achieving pCR was a prognostic factor for OS and PFS.

**Graphic 1.**
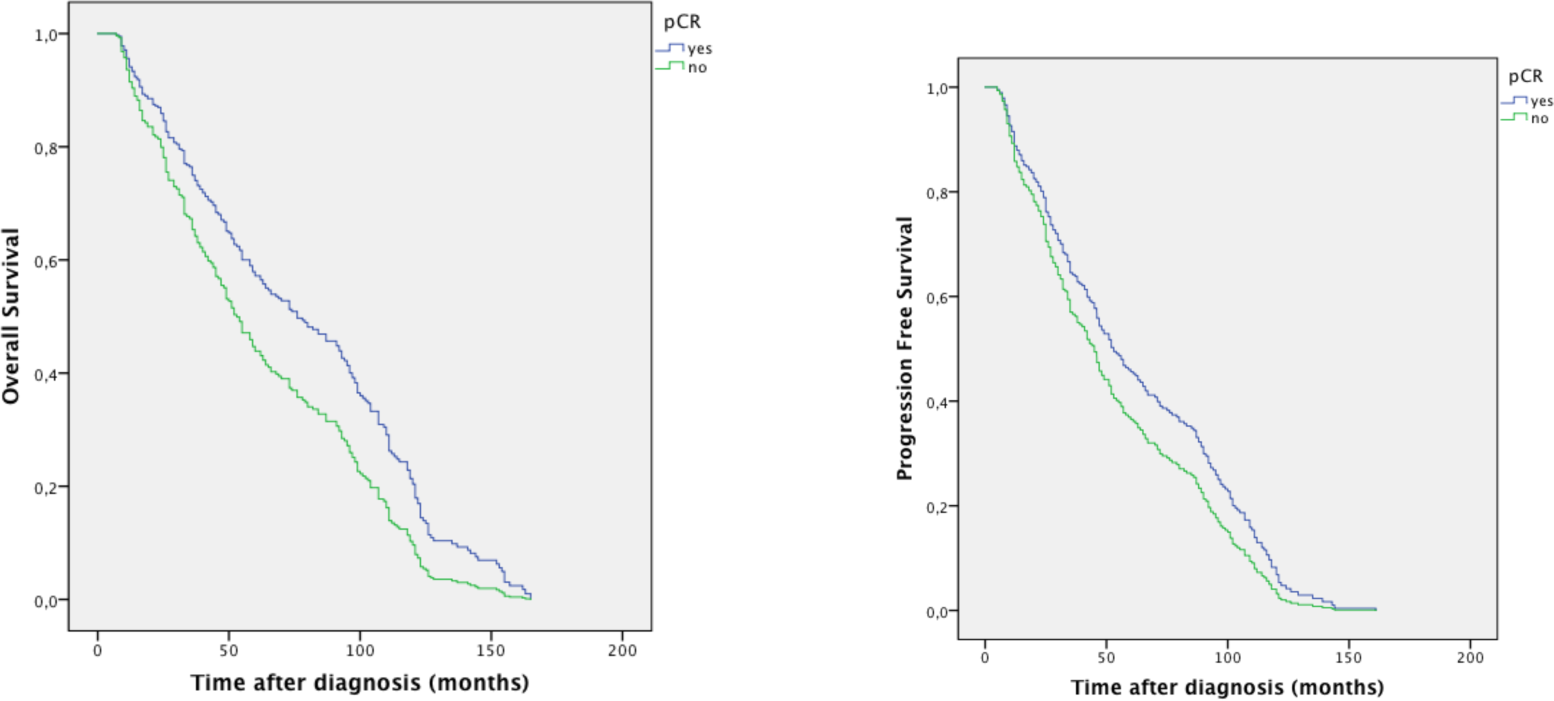
OS and PFS multivariate analysis, based on pCR.

**Graphic 2.**
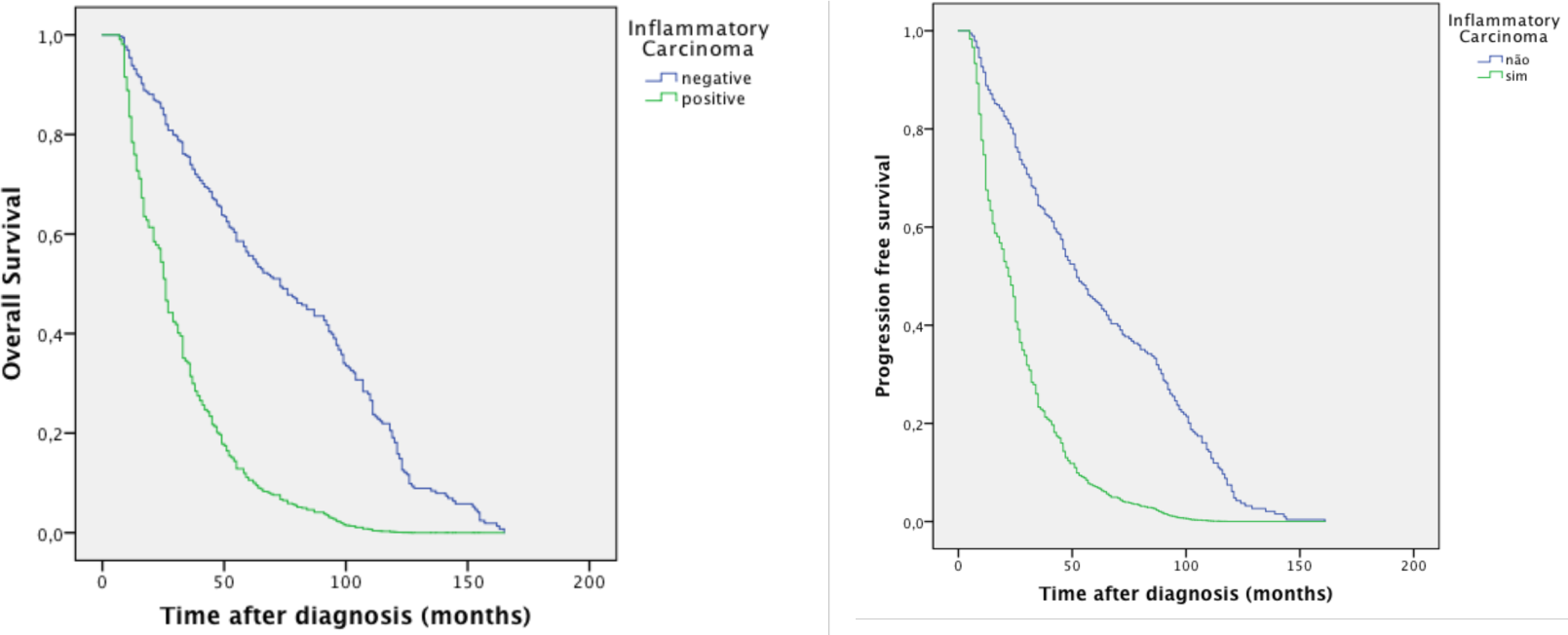
OS and PFS multivariate analysis, based on inflammatory carcinoma.

Multivariate analysis confirmed that patients with pCR had better OS (HR 0,68, CI95% 0,53-0,87, p=0.002) and better PFS (HR 0,78, CI95%, 0,61-1,00, p=0,05).

### Survival analysis according to clinical and pathologic variables

In order to assess which clinical and pathologic variables were prognostic factors for PFS and OS, we performed survival analysis with a median follow-up of 61 months (range 7 - 165 months). During that period, a total of 97 (28,4%) women relapsed and 71 (20,8%) died.

We observed an increased rate of relapse with larger lesions (cT<=2 22,9% vs cT>2 41%, p-value <0,001) and clinically-positive lymph nodes (34,1% vs 19,5%, p-value 0,004), accompanied by increased mortality rate in both groups (cT<=2 14% vs cT>2 36,2%, p-value 0,001; positive nodes 26,9% vs negative nodes 11,3%, p-value 0,001).

Statistically, Patients with inflammatory breast cancer had a significantly worst prognosis in terms of OS and PFS. About 59,1% women with inflammatory carcinoma relapsed during follow-up and 50% died, in contrast to 26,3% and 18,8%, respectively, from general population (p-value <0,001).

Multivariate analysis confirmed that it is associated with worse OS (HR 0,26, CI95% 0,11-0,64, p=0.003) and worse PFS (HR 0,30, CI95%, 0,12-0,74, p=0,009).

In comparison with the impact on pathologic response, increased histologic grade was progressively associated with lower overall 5-year survival (82% G1 vs. 50% G2 vs 38% G3, p-value 0.017) and disease-free survival (64% G1 vs 38% G2 vs 28% G3, p-value 0.040).

By examining molecular subtypes, we verified that Luminal A had the highest relapse rate, but HER2-enriched had the lowest survival rate, as described by Graphic 3.

**Graphic 3.**
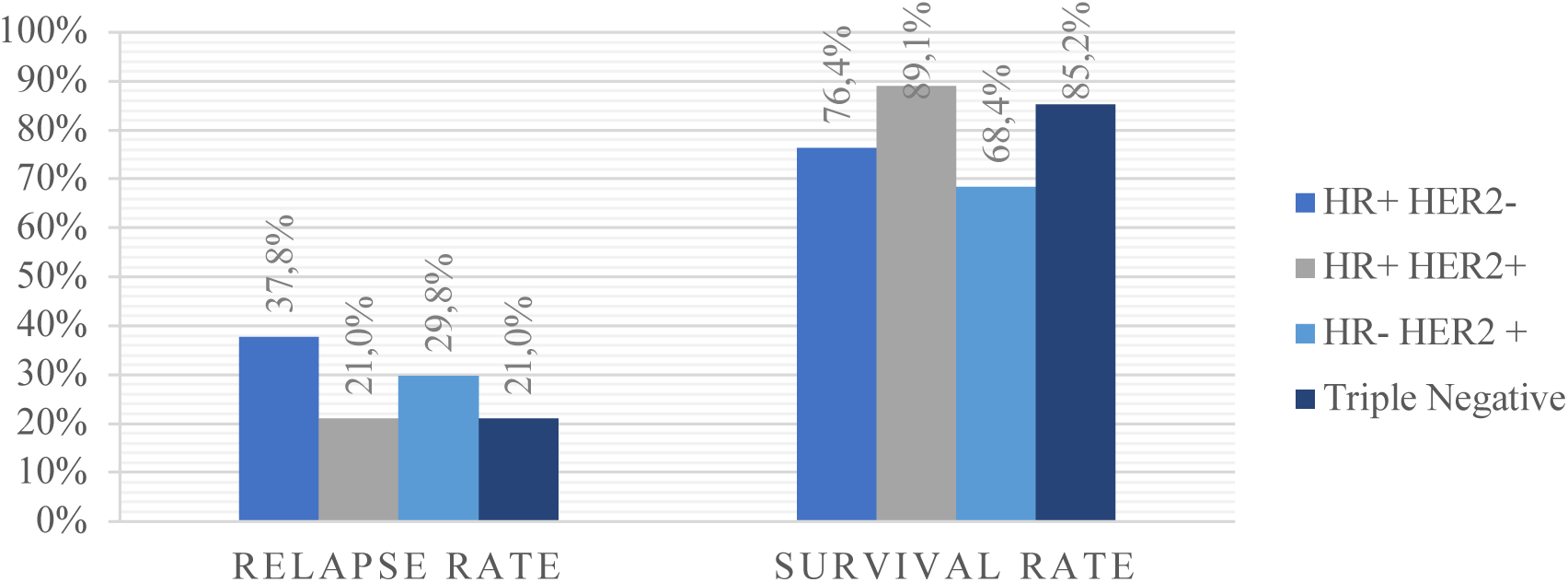
Relapse and survival rate according to molecular subtype (p-value 0,001 and 0,019, respectively)

Regarding molecular markers, we investigated the expression of CK19, CK5 and cadherin E. Although CK19 expression had influenced positively pCR, multivariate analysis demonstrated that its expression was a negative prognostic factor for OS and PFS (Graphic 4).

**Graphic 4.**
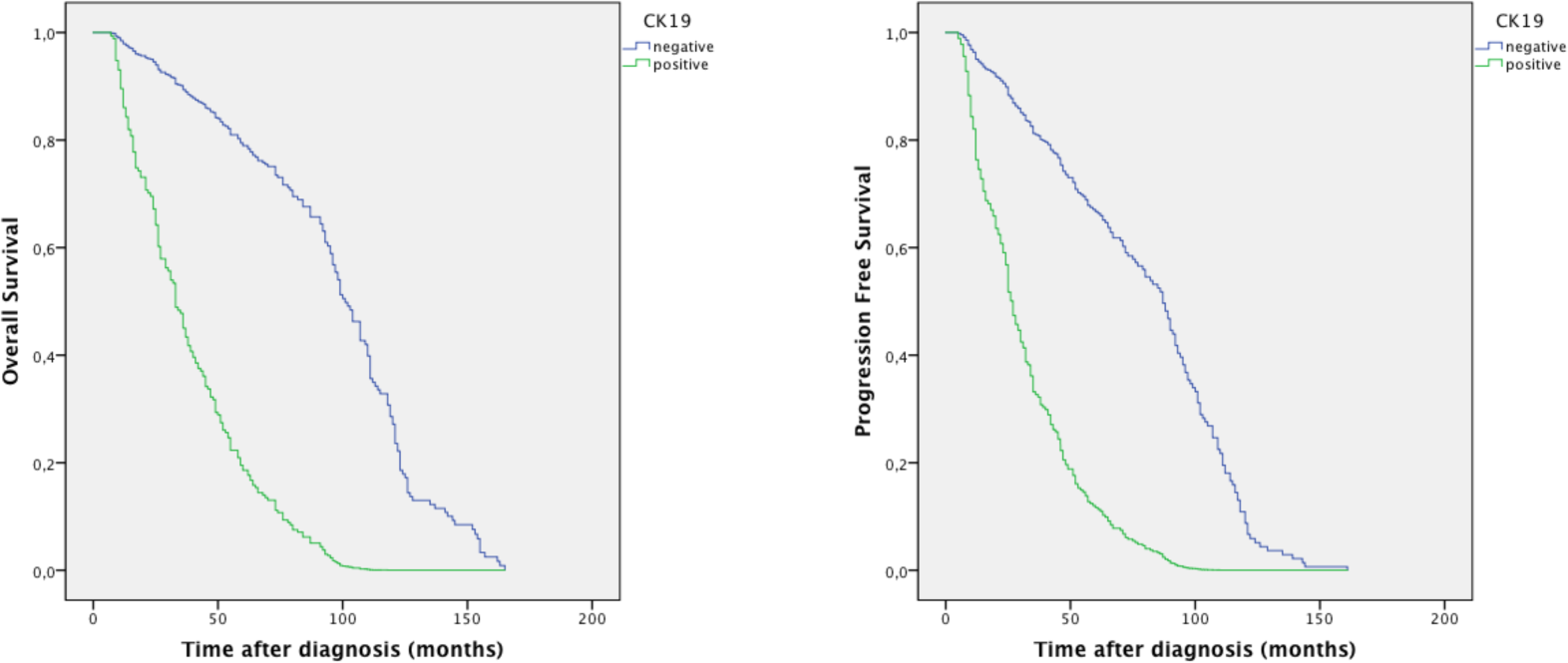
OS and PFS multivariate analysis, based on CK19 expression.

Multivariate analysis confirmed that CK19 expression is associated with worse OS (HR 0,14, CI95% 0,10-0,20, p<0,001) and worse PFS (HR 0,18, CI95%, 0,14-0,26, p<0,001).

By sub-analyzing CK19 population, we determined that its expression was associated with higher histologic grade (75,7% vs 64% grade 3), lower HER2 over-expression (29,3% vs 35,3%) and lower hormone receptor expression (55,7% vs 62,2%).

Younger age, BRCA mutation, multifocality/multicentricity, bilaterality, cadherin E expression and CK5 expression didn’t disclose a significant impact on long-term outcome.

## Discussion

In 1989, Carter et al. described the impact of tumor size, lymphatic extension and clinical TNM staging in patients’ prognosis.^4^ Since then, several other factors were investigated and determined to predict clinical and pathologic outcome, namely tumor biology, tissue or nuclear markers,gene expression profiles and circulating tumor cells.^3,5,6,7,8^ Our study suggests several characteristics that influence prognosis and tumor response to primary chemotherapy.

A complete pathologic response to primary chemotherapy is predictive of better long-term overall and progression-free survival, as was demonstrated by our analysis.^9,10^ CK19 expression, CK5 expression, histologic grade, molecular subtypes and bilaterality behaved as independent predictors for pathologic response.

CK19 is a member of keratin family of proteins, involved in cell signaling, stress response and apoptosis.^11^ It is a target on circulating tumor cell research, as a marker for patients with high relapse risk.^12^ We verified that histologic expression of CK19 in neoplastic breast tissue correlated with higher probability of complete pathologic response and downstaging. However, it was also related to decreased progression free survival as well as overall survival, indicating that CK19 expression has prognostic value in such breast cancer, as described by Kabir et al. (2014).^11^ CK19 population characterization revealed co-existence with other negative prognostic factors, such as higher histologic grade and low expression of hormone receptors, that may have influenced the outcome.

CK5 expression, that is frequently associated with basal-cell phenotype^13^, was equally related to higher probability of pathologic response, but showed no impact on progression free or overall survival.

Although the presence of inflammatory breast cancer didn’t influence pathologic response rate, our study proposes that it negatively influences long-term prognosis, as it increases relapse rate and decreases overall survival.^14,15^ Liu et al. (2017) describes inflammatory carcinoma as a heterogeneous aggressive disease with distinct molecular subtypes associated with different outcomes and chemosensibilities.^16^ However, due to low frequency of patients with this disease, it wasn’t possible to analyze subtypes in this sample.

Histologic grade, molecular subtypes and TNM staging were also prognostic for long-term survival after neoadjuvant chemotherapy.^1,17^

Research is underway to address proliferation and invasion markers, in-depth genomic profiles and circulating tumor cells, that will influence our multidisciplinary approach to locally advanced breast cancer in the future.

## Conclusion

Breast cancer is a complex and heterogeneous disease displaying considerable variation in its clinical and biological characteristics, and thus this cancer remains a challenge of management. Understanding the prognostic and predictive factors for primary chemotherapy will provide an opportunity for a personalized treatment approach.

In summary, this study suggested that cytokeratin 19 expression, cytokeratin 5 expression, histologic grade, molecular subtypes and bilaterality are independent predictors for a complete pathologic response to primary chemotherapy, which influences positively overall survival and progression free survival.

It also established inflammatory carcinoma and cytokeratin 19 expression as independent predictors for worse prognosis.

## Data Availability

The data that support the findings of this study are available on request from the corresponding author, CB.

## Notes

### Competing Interest Statement

The authors have declared no competing interest.

### Funding Statement

No funding was received.

